# Analyzing the Impact of a Real-life Outbreak Simulator on Pandemic Mitigation: an Epidemiological Modeling Study

**DOI:** 10.1101/2022.02.04.22270198

**Authors:** Ivan Specht, Kian Sani, Bryn C. Loftness, Curtis Hoffman, Gabrielle Gionet, Amy Bronson, John Marshall, Craig Decker, Landen Bailey, Tomi Siyanbade, Molly Kemball, Brett E. Pickett, William P. Hanage, Todd Brown, Pardis C. Sabeti, Andrés Colubri

## Abstract

Operation Outbreak (OO), an app-based, educational outbreak simulator, seeks to engage, educate, and empower citizens to prevent and better respond to infectious disease outbreaks. We examine the utility of OO for further understanding and mitigating the spread of communicable diseases. The OO smartphone app uses Bluetooth to spread a virtual pathogen among nearby participants’ devices to simulate an outbreak, providing an experiential learning opportunity. Deployed at many college campuses and other settings, the app collects these anonymized spatio-temporal data, including the time and duration of the contacts among participants of the simulation. Here, we report the distribution, timing, duration, and connectedness of student social contacts at two university deployments and uncover cryptic transmission pathways through individuals’ second-degree contacts. We then construct epidemiological models based on the contact networks generated by OO to predict the transmission pathways of hypothetical pathogens with varying reproductive numbers. Finally, we show that the granularity of OO data enables institutions to improve outbreak mitigation by proactively and strategically testing and/or vaccinating individuals based on individual social interaction levels.

**BIGGER PICTURE:** Outbreak simulation technology can greatly enhance individual and community pandemic preparedness while also helping us understand and mitigate outbreak spread. Building on an existing platform called Operation Outbreak (OO)––an app-based program that spreads a virtual pathogen via Bluetooth among participants’ smartphones––we demonstrate the power of this approach. We investigate the first- and second-degree contacts of OO participants, analyzing the differential risk associated with various local contact network structures. We use OO data to construct an epidemiological model, with which communities may predict the spread of infectious agents and assess the effectiveness of mitigation measures. Based on our findings, we advocate for the wider adoption of outbreak simulation technology to study the implications of social mixing patterns on outbreaks within close-knit communities, to aid pandemic preparedness and response.

## BACKGROUND

Infectious disease outbreaks have repeatedly emphasized the potential for detailed contact tracing data to improve public health [1][2][3]. The COVID-19 pandemic in particular saw the rapid development and deployment of contact tracing technologies in an effort to curb the spread of the virus. However, despite the theoretical benefits of such technologies, adoption rates were often low, stemming from numerous factors including a lack of enforceability and privacy concerns. Without a critical mass of users, these technologies failed to capture the majority of transmission links, compromising their effectiveness [4][5][6]. Moreover, many contact tracing platforms, such as those built upon the Google-Apple Exposure Notification (GAEN) application programming interface (API), generally operated on the principle that contact network data would never be shared unless a user were to test positive [7]. While such a policy benefits the user from a privacy standpoint, it neglects the possible benefit of knowing the user’s typical social patterns and, if needed, intervening accordingly. Finally, the pandemic consistently pointed to young adults in educational settings (e.g., college campuses) as being disproportionately likely to spread COVID-19 [7][8][9]. Yet young adults generally expressed a particular lack of willingness to adopt digital contact tracing technologies [10][11].

To facilitate children and young adults’ engagement in public health, we built an experiential education platform called OO that enables scenario planning for infectious disease outbreaks [12]. OO consists of a suite of tools for learners that includes a smartphone app, a textbook, and a multi-disciplinary curriculum. The smartphone app simulates the spread of a pathogen through a population by transmitting a ‘virtual pathogen’ between participating phones within a threshold proximity. The app also collects anonymous data on the time, duration, and distance of all close contacts between users, as typical contact tracing apps do. OO then processes these simulated transmissions into summary statistics useful for students, teachers, and administrators alike, including levels of social interaction and risk of exposure broken down by participant as well as for the group at large. These statistics also feature as part of the OO curriculum, allowing participants to engage directly with epidemiology through experiential learning.

In this paper, we quantify the social interaction patterns observed among over 750 participants of two OO simulations conducted at two universities in the US: Colorado Mesa University (CMU) and Brigham Young University (BYU). Then, we assess the epidemiological implications of our findings through modeling. We first provide a graphical analysis of the contact networks, focusing in particular on first- and second-degree contacts and the relationship between known and unknown transmission pathways. We also analyze the times and settings that pose the greatest risk for viral transmission. Finally, based on the OO data, we construct an epidemiological model to measure the efficacy of mitigation strategies informed by OO––in particular, diagnostic testing and vaccinations.

## METHODOLOGY

### Simulation Methodology

The OO app, which gathered all data used in this study, is available to the general public on the Apple App Store and the Google Play Store. Upon opening the app, users input a simulation code provided by an OO administrator in order to join a simulation. During the simulation period, the OO app uses Bluetooth Low Energy (BLE) communication to record all proximate interactions between OO participants up to a distance of approximately 3 meters and at a resolution of 1 second. Some of these interactions result in simulated viral transmission if one party is in the infectious state, with the probability of transmission per unit of time being prespecified in the parameters of the simulation. The contact detection over Bluetooth was implemented using a cross-platform software library for iOS and Android called p2pkit [13], which combined public Bluetooth APIs provided by each mobile platform with platform-specific technology, such as WiFi-direct, to maximize proximity sensing. Participants may engage in various ‘interventions’ (e.g., receive virtual masks, PPE, or vaccines) by scanning physical QR codes distributed by the OO administrators throughout the simulation. All events over the course of an OO simulation––contacts, transmissions, use of interventions, recoveries, deaths, and more––are recorded in a backend database that houses the dataset used for this study.

### Recruitment

At CMU, we primarily sought to recruit first- and second-year on-campus students, many of whom had high levels of involvement in on-campus activities and policymaking. This presumably led to some positive bias in their levels of interaction. Our main goal at CMU was to empower students with information on their close contacts and encourage them to consider the epidemiological impact of their social behavior. At BYU, we mainly advertised to individuals studying the life sciences, with the goal of generating data about student behavior. Unlike at CMU, BYU OO participants received daily summary statistics for the simulation including the total numbers of new contacts, infections, recoveries, and deaths. We recruited a total of over 750 participants between CMU and BYU. At CMU, 327 students signed up to participate. The CMU simulation lasted 6 days, between October 29th and November 4th, 2020, which included Halloween weekend. At BYU, 460 participants signed up to participate, comprising both students and BYU faculty. The BYU simulation lasted 9 days, between February 19th until March 1st, 2021.

### Student Engagement

CMU and BYU students expressed an overall willingness to share some of their personal data in order to engage in the outbreak simulation experience. Beyond contributing and analyzing their own data, many students took advantage of the opportunity to learn more about public health. In particular, both CMU and BYU student participants exhibited strong interest in learning how a system for tracking close contacts during an outbreak can help mitigate outbreaks and how individual interactions can disproportionately impact campus-wide health. Across both simulations, however, we observed little change in students’ behavior depending on their epidemiological state within the game, i.e. susceptible, infectious, vaccinated, etc., which likely improved reliability of the social network data but lessened the similarity to an actual outbreak. Overall, the educational focus of OO made it well-positioned to gather data useful to epidemic mitigation without appearing as a threat to students’ privacy.

### Epidemiological Model

Leveraging the anonymous contact networks generated by OO, we propose a method by which such data may be used to construct an epidemiological model that simulates the spread of pathogens and measures the impact of mitigation measures. While the model constructed here does not necessarily reflect any individual institution, we show that the critical assumptions made reflect observations at both CMU and BYU, and as such, the methodology may be applied to either university and likely many others.

The construction of an epidemiological model based on OO data centers around two key inference steps: *network-based inference* and *time-based inference*. While OO reliably captured the time and duration of most interactions between participants, these participants typically represented only a fraction of the institutional population. Moreover, the simulations at CMU and BYU lasted 6 days and 9 days, respectively; epidemiological models for infectious diseases typically require longer time periods to derive meaningful results. We proceeded by applying a combination of parameter estimation and bootstrap techniques to generate the university-wide contact network based on a sample thereof. We then spread a pathogen throughout this network stochastically, and used Monte-Carlo methods to draw conclusions about the inferred propagation of the virus.

For the network-based inference step, we assumed that the true number of contacts, *C*, made by an individual who participated in OO over the simulated period follows a negative binomial distribution [14]. We further assumed that given *C*, the proportion of those contacts who *also* participated in OO, follows a binomial distribution with size parameter *C* and probability parameter *p*. We then solved for the distribution of *C* via maximum likelihood estimation (MLE) given the observed number of contacts per OO participant.

The above framework allows us to generate node degrees for the university contact network but does not provide a characterization of the connectivity between nodes. Based on the CMU and BYU OO simulations, we found strong evidence for proportionate mixing, meaning that the probability of two nodes sharing an edge is proportional to the product of their degrees. To substantiate this claim, we regressed the (binary) existence of an edge between two nodes against the product of their degrees, and found a relatively high *R*^2^ at both universities––0.25 at CMU and 0.20 at BYU. Moreover, proportionately-mixed contact networks based on the OO node degrees mimicked the OO network remarkably well at both universities. In terms of network properties, we focused in particular on the *clustering coefficient*, which is the overall probability that any two contacts of a given person *themselves* had a contact, and the *average shortest path length*, which is the shortest path between a pair of nodes, averaged over all such pairs. At CMU, the modeled clustering coefficient was 0.240 on average (95% CrI: 0.228-0.252) versus 0.272 in the actual network; the modeled average shortest path length was 2.33 on average (95% CrI: 2.29-2.37) versus 2.51 in the actual network. At BYU, the modeled clustering coefficient was 0.184 on average (95% CrI: 0.173-0.195) versus 0.243 in the actual network; the modeled average shortest path length was 2.50 on average (95% CrI: 2.46-2.54) versus 2.69 in the actual network. Based on this finding and a presumed lack of other available information about non-OO participants, we applied a proportionate mixing assumption to the model of the full student body, allowing us to stochastically generate contact networks by assigning each node an expected number of contacts and setting the probability of an edge accordingly.

For the time-based inference step, we implemented a bootstrap method, assuming for simplicity that interactions between any given pair of people are cyclical with a period of 1 week. The model in this paper uses 7 days’ worth of BYU contact data as the bootstrap sampling set; for CMU and other simulations lasting less than one week, weeklong bootstrap samples could be generated by amalgamating one-day bootstrap samples, separating by weekday and weekend. We assumed independence between the total duration of interactions between a pair of nodes and the degrees of those nodes, which was justified by the relatively low observed correlation between these factors of 0.119 (CMU) and 0.051 (BYU).

Under each randomly-generated contact network and bootstrap sample of interaction times, we simulated the spread of a pathogen *in silico* on a network with 6,000 agents sampled from the BYU data. We assumed a single index case, sampled based on node degree, who entered the infectious stage at time 0. Letting *f* be the density function of the virus’ generation interval and letting *I*_*ij*_ be an indicator function of an interaction between infectious individual *i* and susceptible individual *j*, we set the probability of transmission from *i* to *j* equal to:

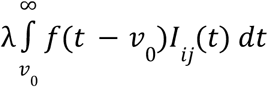

where λ is a constant chosen to reflect the *R*_*e*_ of the virus and *v*_0_ is the time that individual *i* contracts the virus (see [15] for computation of λ; see [16] for comparable methodology). In the event that a transmission occurred (drawn as a Bernoulli trial with probability of success as given in the above equation), we sampled the time of transmission from the density function given by *f*(*t* − *v*_0_)*I*_*ij*_ (*t*), up to a constant of proportionality. As our primary focus in this paper is cumulative cases, and as *f*(*t*) approaches 0 as *t* → ∞, we did not take into account the recovery rate, and assumed reinfection to be negligibly rare.

Finally, we modeled two possible interventions: testing and vaccination. For each intervention, we experimented with a ‘random’ version (in which interventions were administered randomly) and a ‘strategic’ version (in which interventions were administered based on the level of social interaction exhibited per person). We assumed that tests had a constant turnaround time and sensitivity, and that vaccines had already reached a constant and maximum effectiveness level by the start of the simulated period. We further assumed that individuals who test positive would isolate and therefore have no social interactions after the time of receiving the positive result.

We replicated this stochastic model 10,000 times, each time regenerating the node degrees, connectivity matrix, and bootstrap time series samples. We set the model to output the total number of cases at the end of a four-week period. The model was implemented in R, version 4.0.4 [17], with packages igraph [18], lubridate [19], Rfast [20], mixdist [21], and ggplot2 [22].

## RESULTS

We began by investigating OO contact data in order to better understand the differential risk among individuals associated with their contact patterns. First, we simply measured the raw number of contacts per OO participant at both CMU and BYU, filtering out (a) duplicate contacts (multiple contacts between the same pair of individuals); (b) contacts shorter than one minute; and (c) contacts made by persons who did not participate in the entire OO simulation. For the BYU simulation, we only analyzed the first week of data in effort to reduce weekday/weekend bias; for the CMU simulation, we were unable to do so because it lasted only 6 days. Both schools exhibited an overdispersed distribution in the number of contacts per individual, consistent with previous findings (see **Figure 1**, blue distribution) [14]. The mean number of contacts per person was 8.23 at CMU (SD = 11.20, range = 0–58) at CMU and 9.73 at BYU (SD = 13.88, range = 0–82).

**Figure 1:**
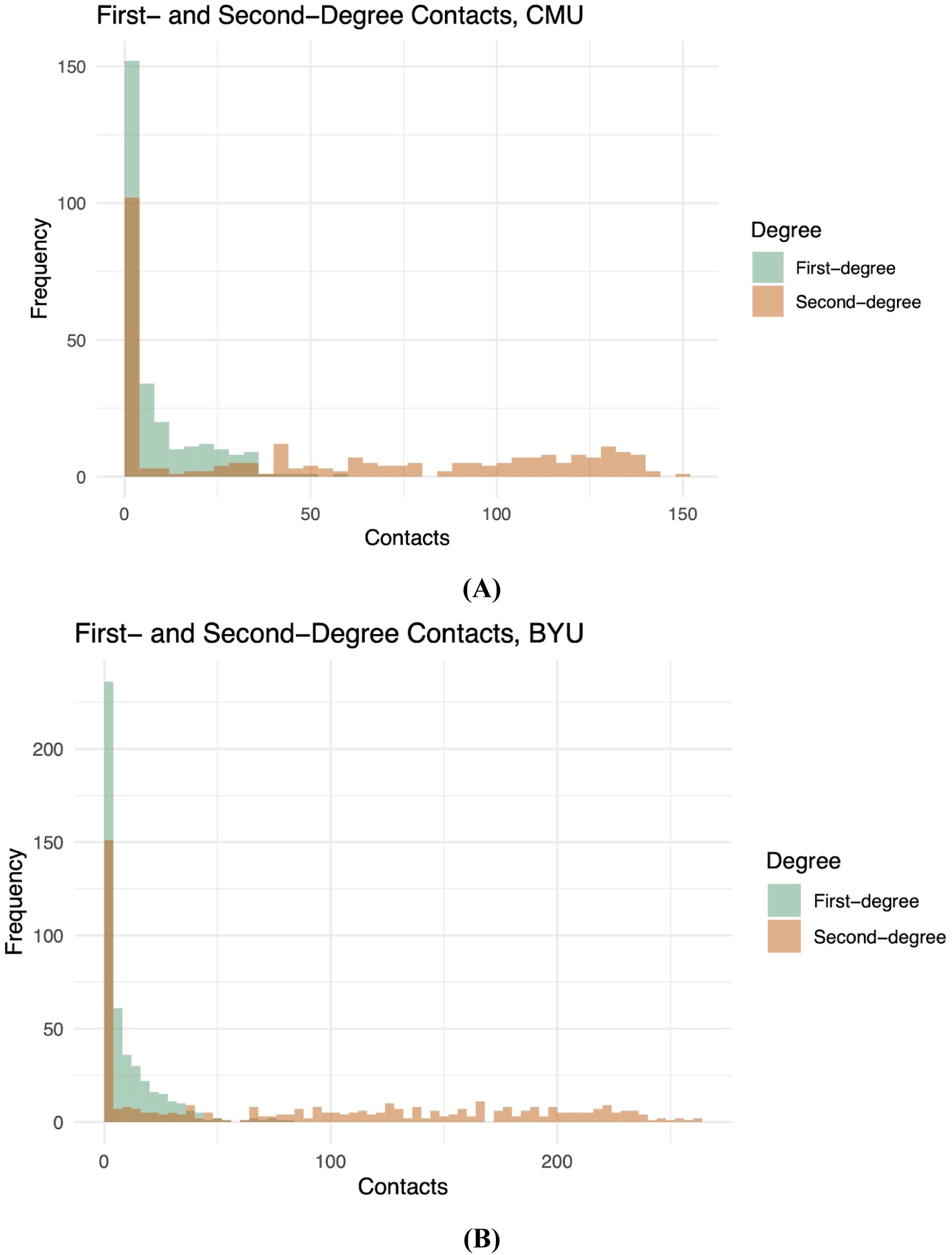
Histogram of contacts per student at **(A)** CMU and **(B)** BYU over the course of one week. MLE for a negative binomial model for first-degree contacts: 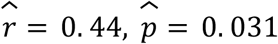 at CMU; 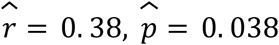 at BYU.

We then looked more closely at the network properties of the contacts. The clustering coefficients––the overall probability that any two contacts of a given person themselves had a contact (see **Epidemiological Model**)––were equal to 0.272 at CMU and 0.243 at BYU. This result is consistent with the findings of Mayer *et al*. (2008) on undergraduate student social network dynamics, which reported a range of 0.17–0.27 for clustering coefficients across 10 American universities based on facebook.com data. To characterize the likely physical environments for these contacts, we also analyzed the time of day/week at which these contacts were most likely to occur, observing spikes during class time at BYU and evenings at CMU. We appreciate that CMU may have exhibited higher-than-normal and otherwise uncharacteristic interaction levels due to social gatherings the night of Friday, October 30^th^, one night prior to Halloween (see **Figure 2**).

**Figure 2:**
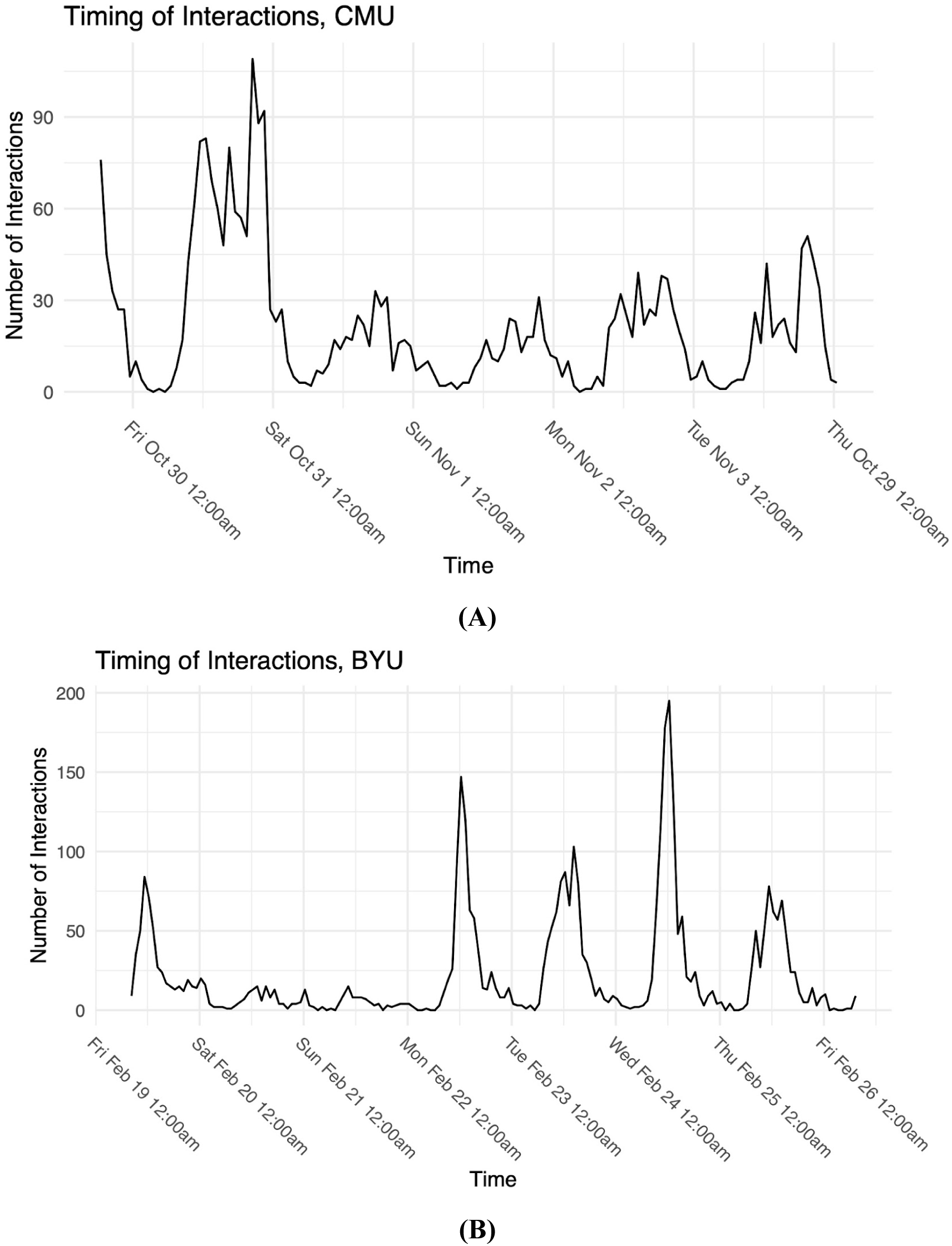
Number of interactions recorded during each hour of simulation at **(A)** CMU among 253 participants and **(B)** BYU amongst 435 participants for which we have complete contact data. The data start on Thursday, October 29th, 2020 at 6:30 pm MDT for CMU and Friday, February 19th, 2021 at 8:18 am MST for BYU. Note that times at CMU *do not* account for daylight savings time, which ended on November 1st, 2020.

We hypothesized that the raw number of first-degree contacts served as a reasonable proxy for risk of infection but could be improved by taking into account (a) durations of contacts and (b) second-degree contacts. Applying the same filtering processes for contacts as described above, we observed high variance in numbers of second-degree contacts for both CMU and BYU participants (see **Figure 1**, red distribution). The mean number of second-degree contacts per person was 53.79 at CMU (SD = 52.30, range = 0–151) and 88.06 at BYU (SD = 85.42, range = 0–264). This analysis gives us a sense of the distribution in the number of second-degree contacts but not the *relationship* between first- and second-degree contacts, which clearly have a strong correlation. As such, we fitted the functional form *y* = α(1 − *e*^−β*x*^) to the number of second-degree contacts as a function of first-degree contacts using least-squares. Despite a relatively low root-mean-square error (RMSE) of 19.1 at CMU and 18.0 at BYU, indicating that the number second-degree contacts can be accurately predicted from the number of first-degree contacts (see **Figure 3**), there were still some individuals whose second-degree contact counts were significantly higher or lower than the model would predict. **Figure 4** presents illustrative examples of an individual who had 7 first-degree contacts but only 32 second-degree contacts, to another with only 3 first-degree contacts but 126 second-degree contacts.

**Figure 3:**
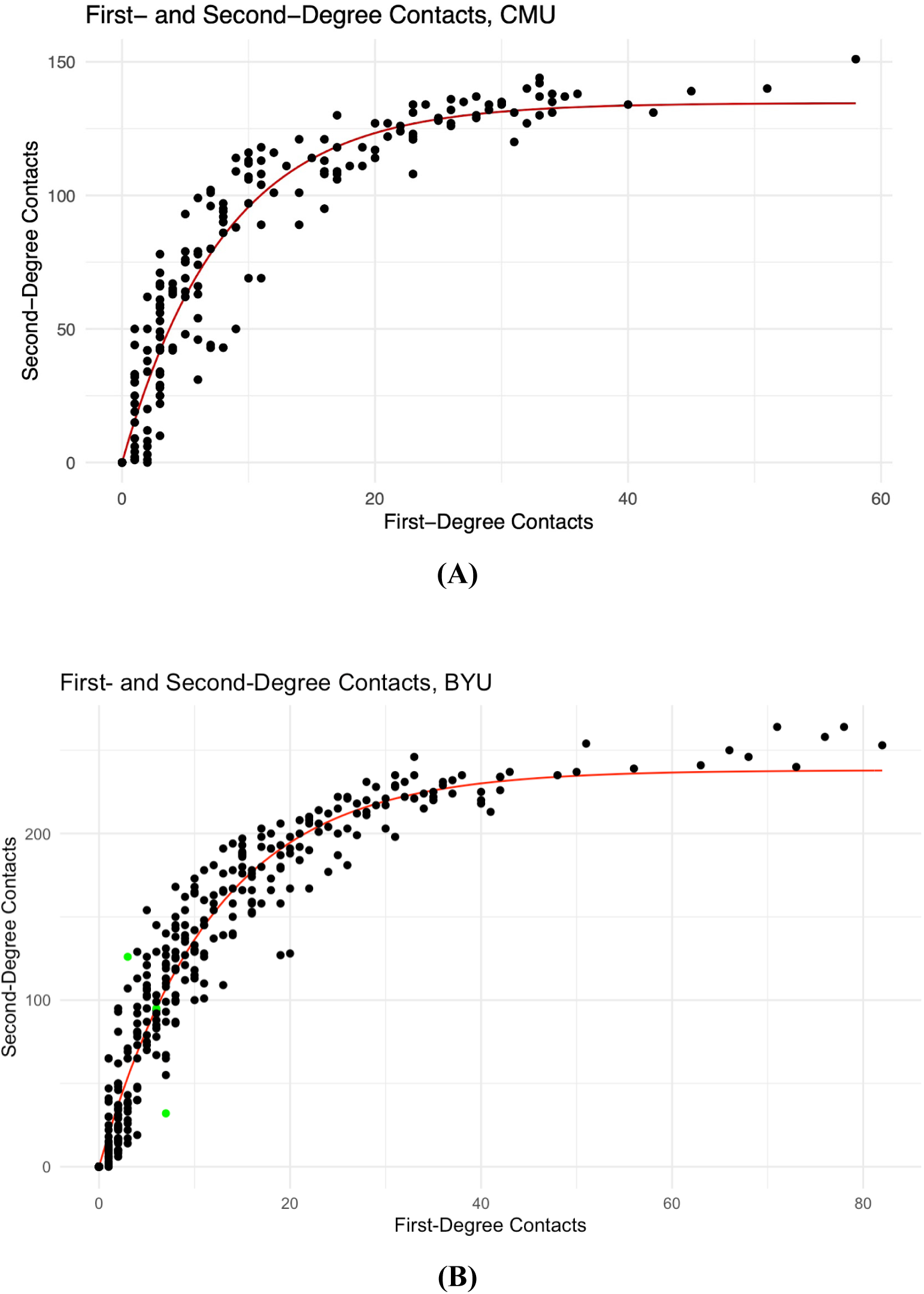
Scatterplot of first degree contacts and second degree contacts for each participant at **(A)** CMU and **(B)** BYU, with the fitted functional form *y* = α(1 − *e*^−β*x*^). Least squares estimates: α = 213, β = 0. 113 for CMU; α = 238, β = 0. 0850 for BYU. The BYU nodes with subgraphs featured in Figure 4 are highlighted in green.

**Figure 4:**
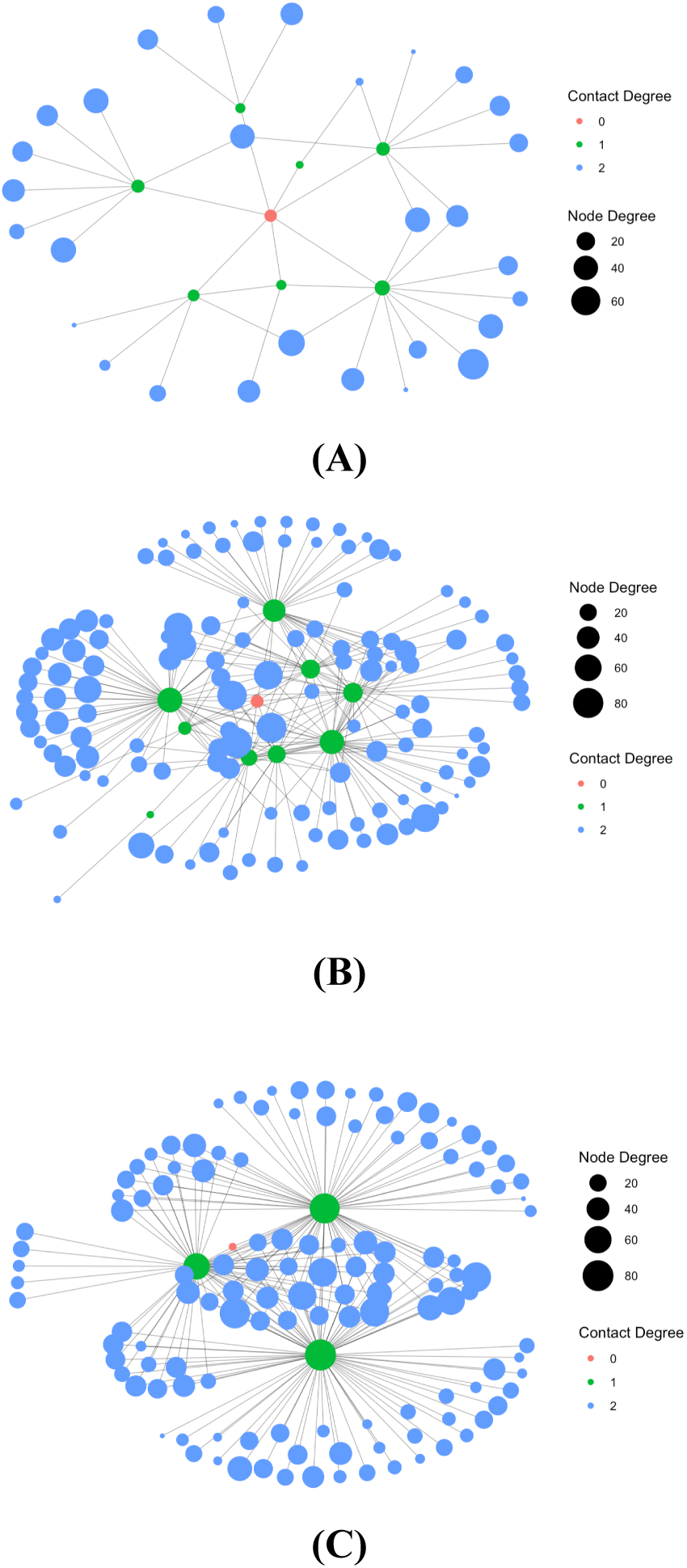
Subgraphs of the BYU contact network in which the number of secondary contacts (blue) is **(A)** lower, **(B)** equal, and **(C)** higher than the model would predict based on the number of first-degree contacts (green). In panel **(A)**, the red node has 7 first-degree contacts but only 32 second-degree contacts;in panel **(B)**, the number of second degree contacts aligns with what we would predict based on the number of first-degree contact; and in panel **(C)**, the red node has only 3 first-degree contacts but 126 second-degree contacts. Edges between second-degree contacts omitted for the sake of visual clarity.

We hypothesized that the *relationship* between first- and second-degree contacts, as well as the durations of such interactions, would leave certain individuals more or less prone to infection than their first-degree contacts alone would suggest. To test this hypothesis, we first simulated the spread of COVID-19 through the real OO contact networks using mean-field approximation, a computationally-efficient method for estimating the probabilities of each person being in each epidemiological state (susceptible, exposed, infectious, recovered) at a given time [23][24]. We then regressed the probability that each individual had been infected against various statistics describing social contacts. We began with two extremely simple statistics: equal-weighted and duration-weighted numbers of contacts. “Equal-weighted” means the number of contacts for an individual; “duration-weighted” means the sum of durations of all contacts for an individual. Assuming no intercept term, the regressions yielded an adjusted *R*^2^ of 0.554 and 0.925, respectively, for CMU and 0.434 and 0.880, respectively, for BYU (see **Figure 5**). These results emphasized the impact of including contact duration in risk assessment.

**Figure 5:**
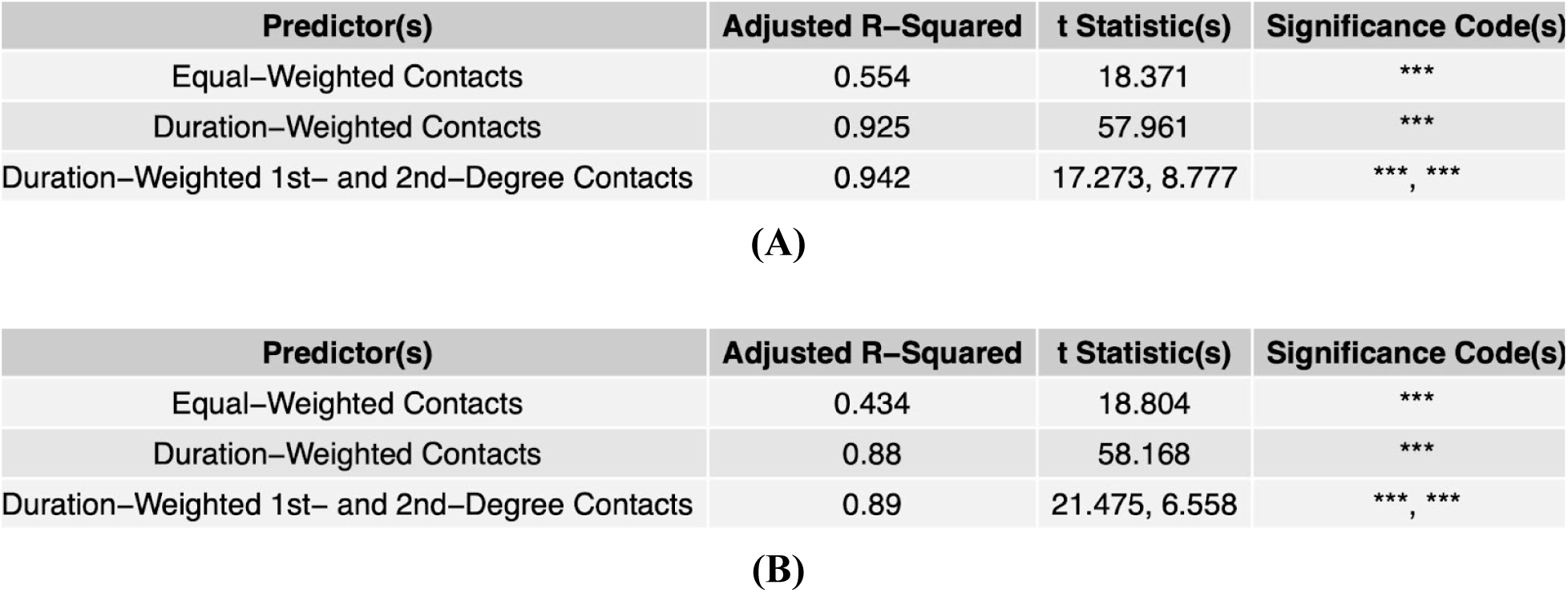
Regression analyses for **(A)** CMU and **(B)** BYU. We modeled the probability of infection at the end of the OO simulation as a linear combination of various factors, including equal-weighted contacts, time-weighted contacts, and time-weighted second-degree contacts.

We found that second-degree contacts made a statistically-significant impact on probability of infection, even beyond what could be captured by first-degree contacts alone. Taking into consideration our previous finding about contact duration, we constructed an additional predictor variable––duration-weighted second-degree contacts––by multiplying the total durations of the two contacts involved. For example: If persons A and B interact for a total duration of 60 seconds, and persons B and C interact for a total of 80 seconds, then the second-degree contact between persons A and C via person B contributes a factor of 4800 to person A’s duration-weighted second-degree contacts. To compute the total value of this statistic for person A, we simply sum over all possible second-degree contacts for person A, including second-degree contacts that are also first-degree contacts. Using duration-weighted second-degree contacts as an additional predictor variable in our regression analysis, we found high statistical significance, as well as a slight increase in adjusted *R*^2^ as compared with duration-weighted first-degree contacts alone (see **Figure 5**).

We first ran the epidemiological model varying only the basic reproductive number (R_0_) to reflect differences in infectivity associated with different variants of COVID-19. Using kernel density estimation from the Monte-Carlo simulation, we determined the distribution of the cluster size after a four-week period assuming one initial case. Under all simulations, this distribution was positively skewed––increasingly so with higher values of R_0_. See **Figure 6** for a summary of the results.

**Figure 6:**
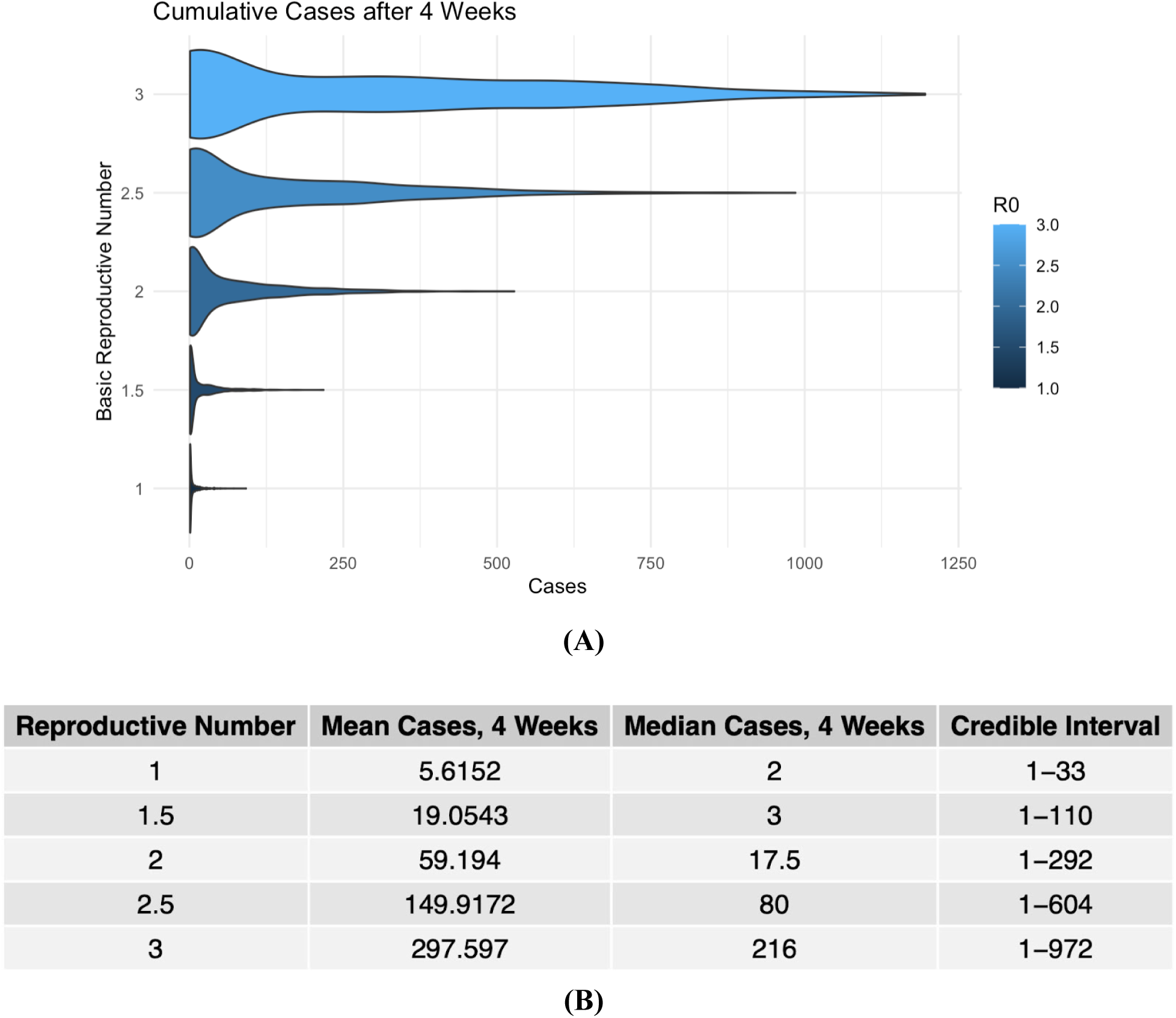
Results of the epidemiological model under five different possible values of R_0_ ranging from 1 to 3. **(A)** Results expressed as a density estimate; **(B)** summary statistics from each model run.

We then measured the impact of diagnostic testing and vaccinations, which could be implemented according to a random strategy (*i*.*e*., equal probability of testing/vaccination for everyone) or an OO-based strategy (*i*.*e*., probability of testing/vaccination proportional to social activity level). Under four different levels of testing and vaccination, the OO-based strategy drastically reduced the reproductive number and case counts, with a smaller number of tests/proportion vaccinated corresponding to a more dramatic reduction (see **Figures 7-8**).

**Figure 7:**
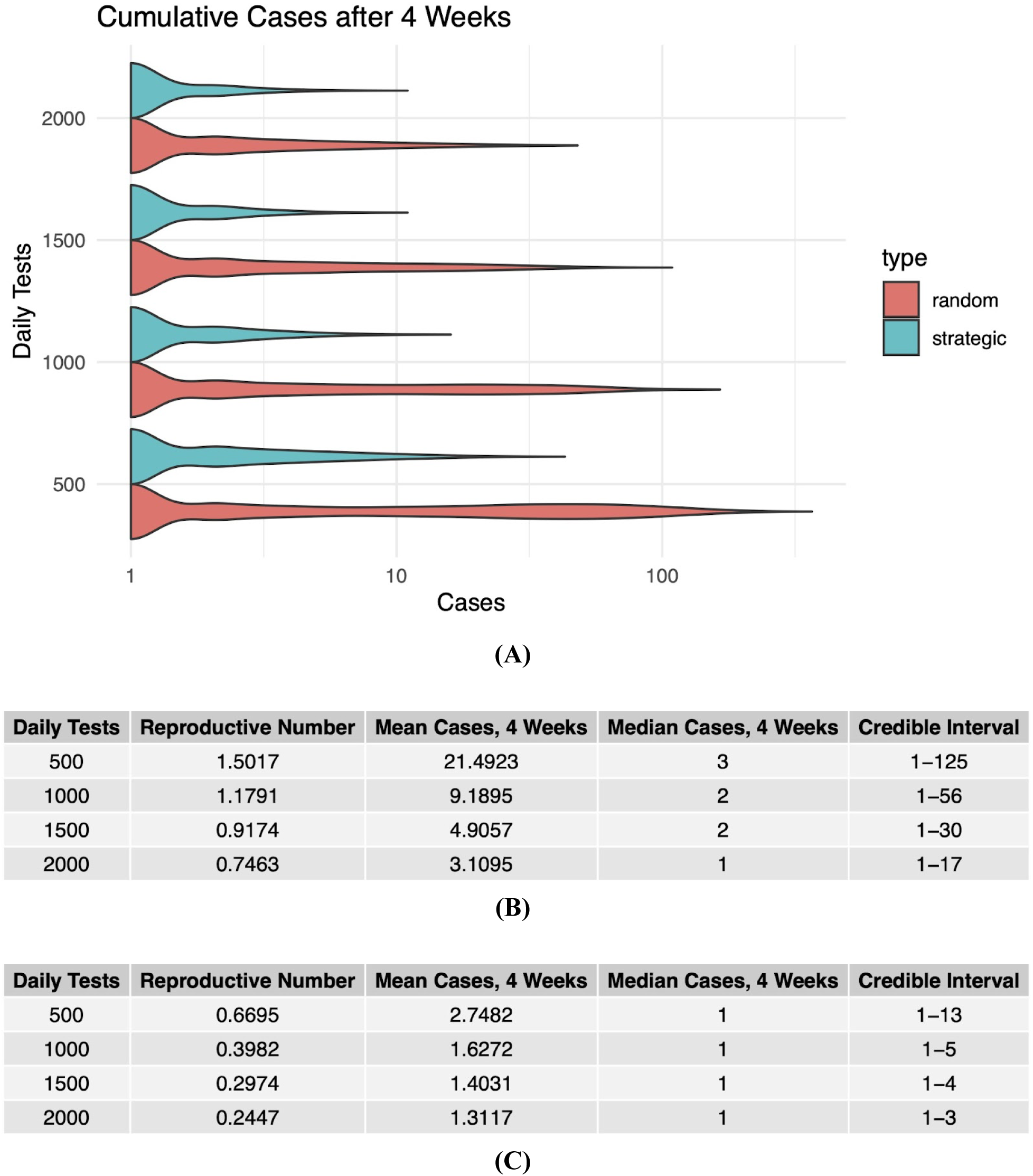
Results of the epidemiological model under four different possible testing rates, ranging from 500 to 2000 per day, administered either randomly or based on activity level. **(A)** Results expressed as a density estimate; **(B)** summary statistics from each model run under random testing; **(C)** summary statistics from each model run under strategic testing.

**Figure 8:**
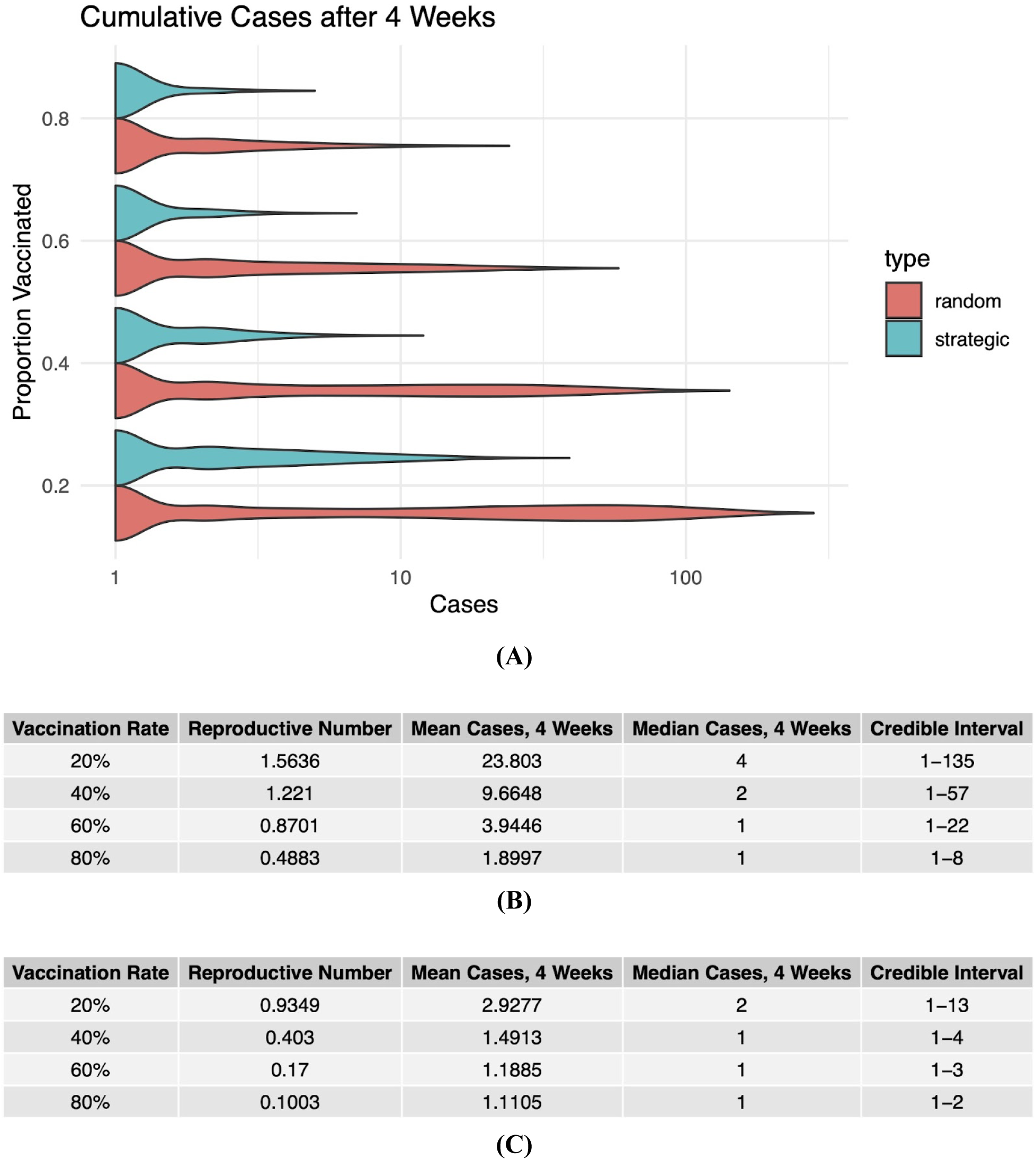
Results of the epidemiological model under four different possible vaccination rates, ranging from 20% to 80%, administered either randomly or based on activity level. **(A)** Results expressed as a density estimate; **(B)** summary statistics from each model run under random vaccination; and **(C)** summary statistics from each model run under strategic vaccination.

## DISCUSSION

The OO data we gathered offer a number of substantive conclusions about how close-knit communities such as schools and universities should factor social interaction patterns into their pandemic response. Beyond providing a distribution of the volume, duration, and timing of social contacts, a deeper look at the OO contact network structure reveals the added risk of *cryptic* transmission pathways––that is, pathways largely unbeknownst to the infectee as a result of the variance in the distribution of second-degree contacts. As revealed by our regression analysis, these second-degree contacts significantly impact individual-level risk and, as such, may help public health authorities best identify the individuals who are most liable to contracting or transmitting the virus.

We then propose a framework by which OO-participating institutions may construct an epidemiological model based on OO network data. Notably, these models rely on statistical inference techniques that allow them to be constructed even if only a fraction of institution members participate in OO. Based on these models, institutions may view how various pathogens with different epidemiological parameters will likely propagate through the population.

We demonstrate the potential benefit of using OO social activity data as a means of strategically testing and/or vaccinating individuals in a population. While such a strategy hinges on a high OO participation rate relative to the population (which we observed neither at CMU nor at BYU), the theoretical reduction in cumulative cases is drastic, even under relatively low levels of testing and/or vaccination. Any such proactive risk-based measures, however, would have to be implemented thoughtfully so as not to incentivize riskier behaviors. This is a place where such an educational outbreak simulation can further be useful, as an opportunity for communities to think through varying behavioral responses and outcomes in a low-stakes setting.

The data generated by the OO app may be used for further epidemiological analyses of various SARS-CoV-2 transmission characteristics, such as the high overdispersion that results in most introductions going extinct. We plan to use the OO networks to investigate the effects of overdispersion on the outbreak dynamics by introducing pathogens with varying levels of this parameter. Our data can also be helpful in distinguishing between virological and behavioral superspreading. In the former, a subset of infections are more infectious per contact, while in the latter all individuals are highly infectious at some point, but the superspreaders happen to make more contacts. We hope such further research will stress the importance of analyzing individual behaviors in the context of infectious disease outbreaks.

## LIMITATIONS

OO comes with some limitations in terms of its reliability of modeling individual-level risk and outbreaks more generally. For example, student interaction patterns may change dramatically between the time of the OO simulation and the time of an actual epidemic. In these particular cases, we ran the simulations during the ongoing COVID-19 pandemic, so the recorded OO data may not reflect typical student behavior. OO simulations can be also run with increased levels of participant engagement, including as a fully immersive outbreak experience. The bluetooth sensing is also blind to the type of interaction, beyond distance, and whether individuals are masked or not. Moreover, assuming a participation rate of less than 100%, there will always be individuals whose social activity levels cannot be computed, so any strategic testing/vaccination plan cannot be tailored to that missing fraction of the population. From a technological standpoint, while Bluetooth-based proximity sensing is widely available on most smartphones, mobile operating systems often pose restrictions to the use of such capabilities. That being said, the recent availability of open-source Bluetooth libraries such as Herald, which contains the basis for the contact tracing app TraceTogether, offers an ongoing solution to research on proximity sensing technologies such as OO.

## CONCLUSIONS

We are more connected than we may think. Social contact patterns observed in the university setting revealed significant variation in local contact networks between individuals, leaving some overexposed or underexposed to risk in ways that the individual may not recognize. Moreover, knowledge of individual-level risk can make a drastic impact on the ability of an institution to mitigate an epidemic. To prepare for the next pandemic, it is essential that we gather social contact data in times of health in order to prepare for times of sickness. Furthermore, a platform such as OO that integrates pandemic education with preparation and mitigation can engage at-risk populations, such as students, and incentivize them to comply with public health interventions by allowing them to be active and informed participants in pandemic response.

## Data Availability

All code and data necessary to run the model and generate the figures is available at https://github.com/broadinstitute/OO-modeling.

## ACKNOWLEDGEMENTS

We would like to acknowledge Fathom Information Design for providing data visualization support and Fuzz Productions for developing the version of the OO app used in the CMU and BYU simulations. We would like to thank our interns at the Broad Institute for their contributions to the OO curriculum, student leaders at CMU and BYU for their coordination of the simulations, and of course, all of our participants.

## CODE AND DATA AVAILABILITY

All code and data necessary to run the model and generate the figures is available at https://github.com/broadinstitute/OO-modeling. The OO datasets were anonymized by assigning each participant an ID number, as well as a corresponding P2P identifier code.

## ETHICS STATEMENT

The Brigham Young University Institutional Review Board determined this work to be non-human subjects research (Protocol IRB2021-207). Harvard Longwood Campus Institutional Review Board of Harvard University determined this work to be non-human subjects research (as is typical for non-human subjects research projects at Harvard, no protocol number was issued). The University of Massachusetts Chan Medical School Institutional Review Board determined this work to be non-human subjects research (Protocol H00024280). The Colorado Mesa University Institutional Review Board determined this work to be non-human subjects research (Protocol 21-44). Lastly, The Broad Institute of MIT and Harvard’s Office of Research Subject Protections determined this work to be exempt (Protocol EX-6991).

## SUPPLEMENTARY MATERIALS

### SUPPLEMENT A: MODEL PARAMETERS

**Figure S1:**
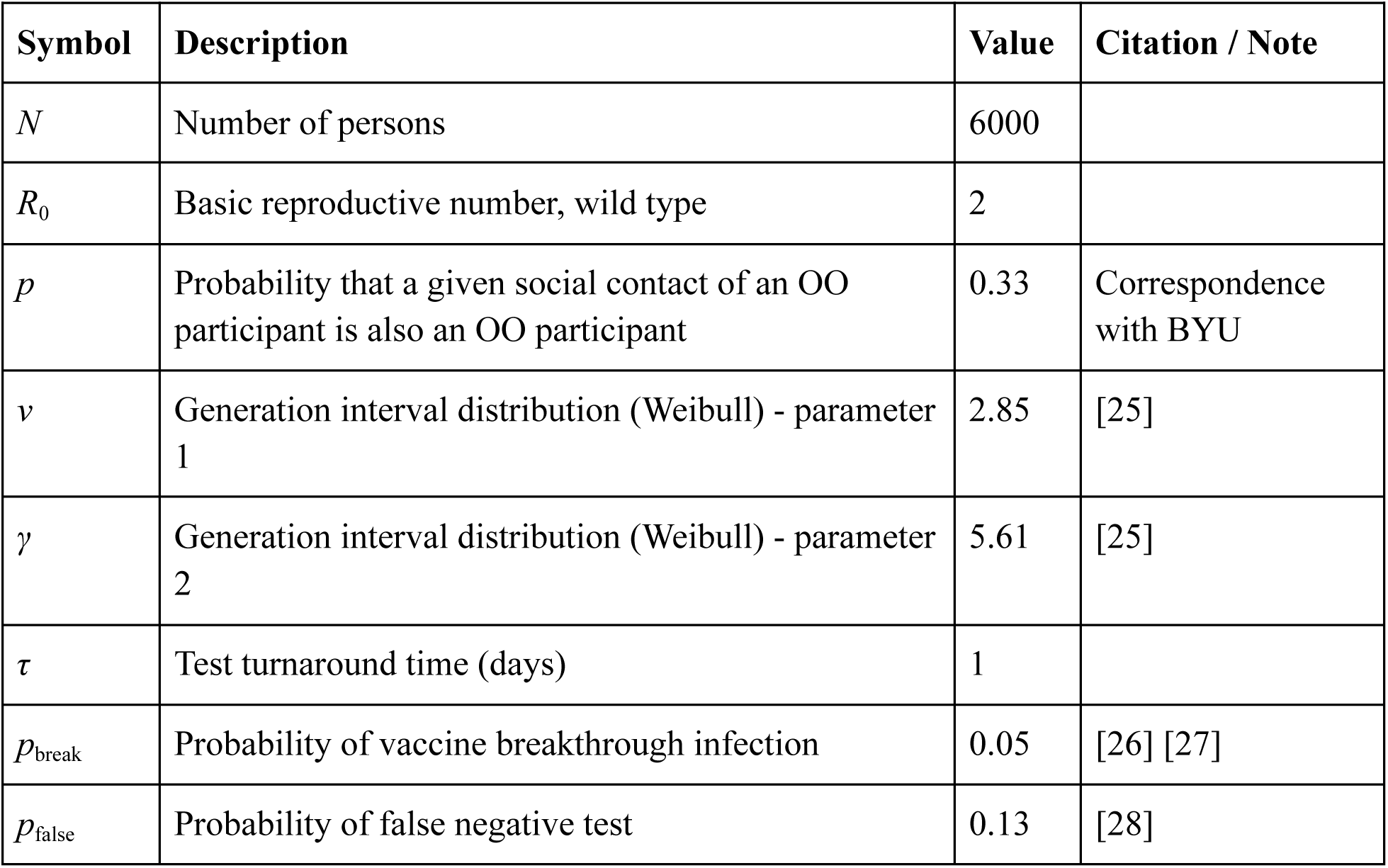
Baseline model parameters

### SUPPLEMENT B: ADDITIONAL RECRUITING INFORMATION

At CMU, we recruited OO participants through flyers, email and social media outreach, prizes for those involved, and word of mouth from student leaders and residence hall assistants. We recruited student on-campus administrators for OO from a pandemic leadership university capstone course. During the simulation, OO student leaders publicized upcoming in-app and out-of-app public health quizzes, answered questions, and announced updates with the participants via a free group messaging platform. Students could respond to the OO leaders directly via the messaging platform to ask for assistance, clarification, or answer out-of-app quizzes to earn in-app benefits such as PPE, tests, and masks through QR codes. We encouraged students to individually review their contacts each day over the six day simulation.

At BYU, the main goal of OO was to generate data about student behavior for a post-simulation faculty seminar to discuss findings. We advertised OO to the student body through various forms of messaging including physical and digital posters, physical and digital newsletters, and a website. We recruited on-campus student administrators from a student-led academic association.

The experiential learning opportunity was offered in various courses, with a subset of professors offering participation as extra credit. We sent information about accessing interventions in the context of the OO simulation (e.g. tests, vaccines) via email.

At both universities, we informed participants of the goals of the simulation and provided assurance that location and other private data would not be collected, but that location-absent anonymous proximity data would be collected. We instructed participants to go about their normal daily routine while adhering to existing real-world COVID-19 policies (e.g., mask-wearing and physical distancing where necessary). Students facilitating the events used lighthearted, positive messaging in their daily updates in order to engage as many of the participants as possible.

